# How Well Do We Do Social Distancing?

**DOI:** 10.1101/2023.02.16.23286061

**Authors:** Naohide Yamamoto, Mia Nightingale

## Abstract

During the pandemic of coronavirus disease 2019 (COVID-19), many jurisdictions around the world introduced a ‘social distance’ rule under which people are instructed to keep a certain distance from others. Generally, this rule is implemented simply by telling people how many metres or feet of separation should be kept, without giving them precise instructions as to how the specified distance can be measured. Consequently, the rule is effective only to the extent that people are able to gauge this distance through their space perception. To examine the effectiveness of the rule from this point of view, the present study empirically investigated how much distance people would leave from another person when they relied on their perception of this distance. Participants (*N* = 153) were asked to stand exactly 1.5-m away from a researcher, and resultant interpersonal distances showed that while their mean was close to the correct 1.5-m distance, they exhibited large individual differences. These results suggest that a number of people would not stay sufficiently away from others even when they intend to do proper social distancing. Given this outcome, it is suggested that official health advice include measures that compensate for this tendency.

---

During the pandemic of coronavirus disease 2019 (COVID-19), several measures have been introduced for reducing person-to-person transmission of the virus that causes the disease (severe acute respiratory syndrome coronavirus 2; SARS-CoV-2). Among them is the so-called social distancing (also known as physical distancing) in which people are asked to maintain a certain distance from each other so as to minimise close contact between individuals who are infected by the virus and those who are uninfected. As of February 2023, the World Health Organization recommends having an interpersonal gap of at least 1 m on the basis of observational evidence that this extent of separation can help reduce the propagation of the virus (Chu et al., 2020; World Health Organization, 2022). However, it is still unclear whether the social distance of 1 m is sufficient because it has been shown that expiratory droplets and aerosols can carry the virus over a longer distance (Guo et al., 2020; Jones et al., 2020).

In addition, there is another factor that can call the effectiveness of social distancing into question: The current approach rests on the (probably implicit) assumption that when people are verbally given an interpersonal distance to maintain, they can use this information to perceive and adjust the distance accurately. For example, official recommendations for social distancing tend to simply state the person-to-person distance to be kept, or they often come with vague instructions about how to achieve the desired separation (e.g., ‘two big steps’; Queensland Government, 2022a). Similarly, typical signage for social distancing just contains verbal descriptions of the recommended distance (such as ‘keep 1.5 m from others’) without indicating the distance physically (Figure 1). When implemented this way, the social distancing measure is effective only to the extent people are able to make an accurate judgement of the interpersonal distance through their space perception ability. To evaluate the effectiveness of the current implementation of the social distancing measure from a perceptual point of view, the present study empirically tested how well people would stay away from another person by using their perception and knowledge of the to-be-given gap alone.

**Figure 1.**
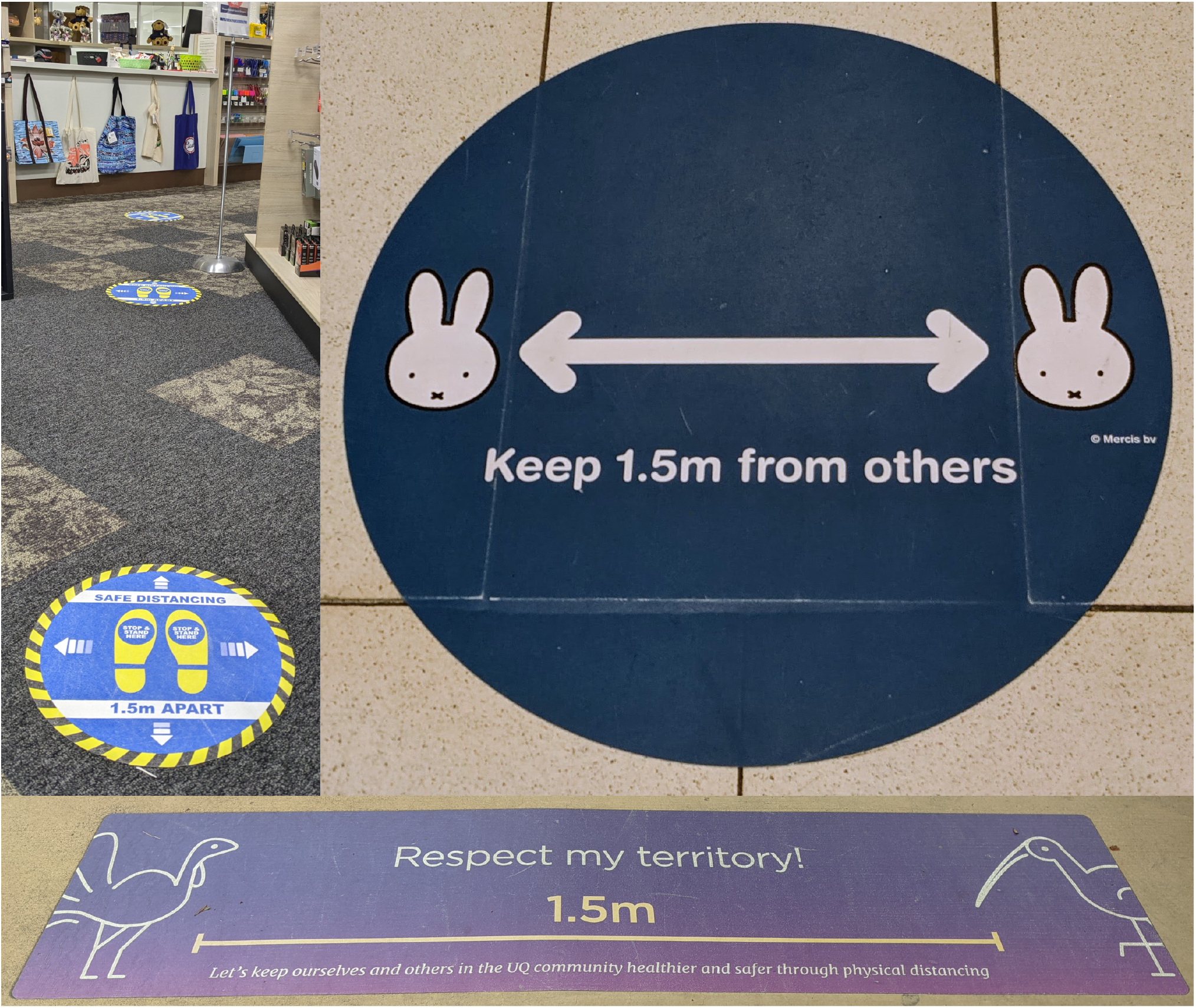
Examples of Typical Signage for Social Distancing. *Note*. All of these signs contain verbal descriptions of the social distance (1.5 m in these cases), but only the bottom one physically indicates the extent of the recommended separation (the horizontal line). Signage can even be misleading (e.g., the top left example may be construed such that the physical distance between two adjacent marks corresponds to the verbally indicated distance, but it does not).

Research on distance perception has shown that people are reasonably accurate in perceiving short distances from themselves to an external object (egocentric distances). This is particularly the case when healthy adults indicate their perceived distance through action (e.g., walking without vision to a briefly previewed target; Loomis & Philbeck, 2008). However, when they use verbal information to judge the target distance, they tend to underestimate it (e.g., saying that a target is 80-cm away when its physical distance is 1 m; Andre & Rogers, 2006; Foley et al., 2004; Yamamoto et al., 2014). This underestimation tendency may actually be beneficial in the context of social distancing because it should make people move farther away—that is, when they are physically at a 1-m distance from another person, they think it is about 80 cm, which should prompt them to increase the interpersonal gap. Thus, it was predicted that when people were verbally asked to leave a given distance between themselves and another person, they would underperceive the physical distance, which in turn should lead them to overproduce the required gap (e.g., leaving 1.8 m when attempting to stay 1.5-m away).

## Method

This study was approved by the Office of Research Ethics and Integrity at Queensland University of Technology (approval number: 4684). It was carried out in accordance with the National Statement on Ethical Conduct in Human Research (National Health and Medical Research Council, 2018).

## Participants

One hundred and fifty-three participants were recruited from students and staff of Queensland University of Technology and also from the local community. They consisted of 64 male, 88 female, and 1 non-binary participants, ranging from teenagers to those in their eighties (Table 1). They volunteered for the study without receiving any incentives. All participants verbally gave their informed consent prior to participation in the study.

**Table 1.**
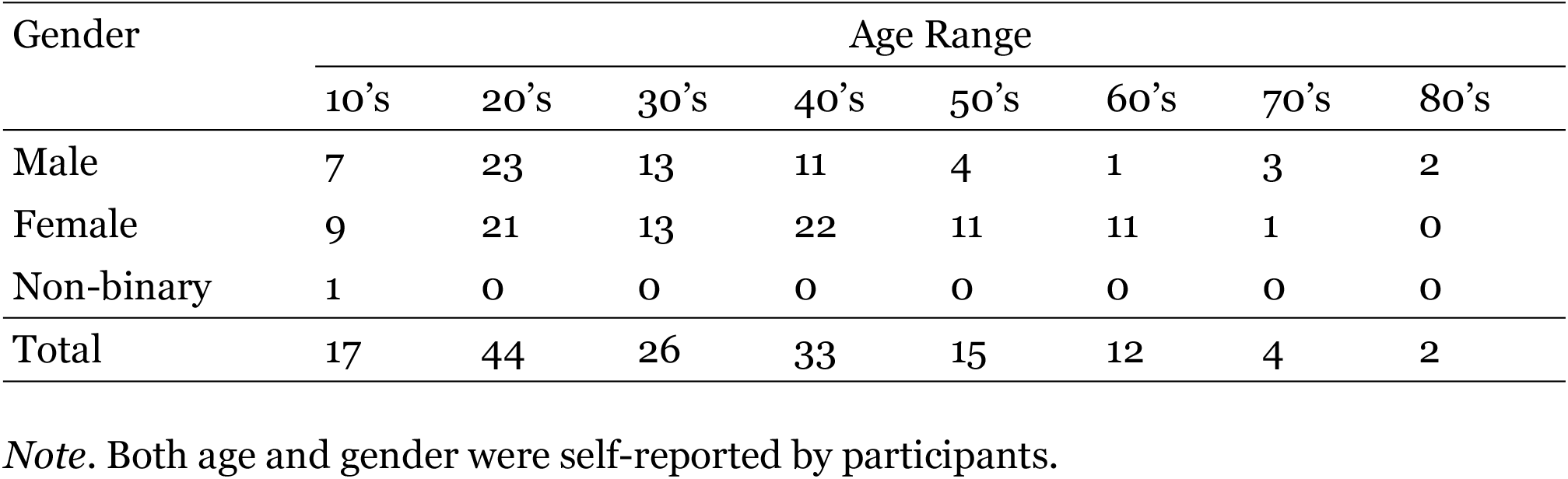
Numbers of Participants per Age Range and Gender.

A priori power analysis showed that a sample of nine participants could have been adequate for finding the predicted overproduction of an interpersonal gap (see Supplementary Material for details). However, such a sample would not have given enough room for including participants of various ages. Thus, the sample size was not set through the power analysis, but instead we aimed to recruit as many and diverse participants as possible to enhance the representativeness of results from the present study.

## Design and Procedure

Data were collected between October 2021 and January 2023 in Brisbane, Australia, where the recommended social distance was 1.5 m. Although situations of the COVID-19 pandemic kept changing (and so did implementation of various preventive measures), the official recommendation for maintaining a 1.5-m interpersonal gap was consistently in place during the data collection period (Queensland Government, 2022b). While this study aimed to test diverse participants, only people in Brisbane were recruited so that all participants were familiar with the same 1.5-m social distance.

Participants were tested individually. At the beginning of testing, a researcher and a participant stood face-to-face. The researcher remained in this initial position and verbally asked the participant to leave exactly 1.5 m between them. The participant was free to view anywhere and move in any manner while adjusting the interpersonal gap. However, the participant was not given any particular information that could aid in gauging the physical distance. Once the participant stopped moving, the toe-to-toe distance between the participant and the researcher was measured using a tape measure placed on the ground. Each participant’s self-reported age (in an approximate form—i.e., being in their 10’s, 20’s, etc.) and gender were also recorded.

The present study aimed to capture social distancing behaviour in an ecologically valid fashion. To this end, participants were tested in a variety of settings so that data would not be biased toward any particular social distancing situations. They included rooms and corridors of office buildings, various outdoor locations in built environments (e.g., footpaths and squares), and open spaces in the park. These testing sites were large, devoid of any obvious distance cues (e.g., no patterns on the floor), and free of obstacles so that the participants were unconstrained while moving closer to and farther away from a stationary researcher.

During the informed consent process, participants were told that this study was to investigate how people perceive distance in naturalistic settings. That is, they were not informed that the study was about social distancing. This was to minimise potential response bias in the present study. In social distancing, leaving too little distance is problematic, while keeping a larger-than-required gap is fine. Thus, if the participants had been explicitly asked to do social distancing, it could have prompted them to overproduce the interpersonal distance. For the same reason, each participant was tested just once. Although it was never made explicit to the participants that the study was to measure social distancing performance, repeatedly performing the task would have provided more opportunities to infer its relation to the 1.5-m social distance. The participants were debriefed about the true purpose of the study at the completion of their participation.

## Results

Original data collected in this study are available on Open Science Framework at https://osf.io/zs7jv/. Figure 2 displays interpersonal distances produced by participants. While the participants made a wide range of responses (0.67–2.94 m), their mean was close to the correct social distance of 1.5 m (*M* = 1.444 m, *SD* = 0.319 m). However, the group mean was still significantly different from 1.5 m, *t*(152) = −2.150, *p* = .033, *d* = 0.174, 95% CI [1.393, 1.495], showing that the participants as a whole underproduced the required distance. Indeed, 61.4% of the participants (*n* = 94) produced distances shorter than 1.5 m.

**Figure 2.**
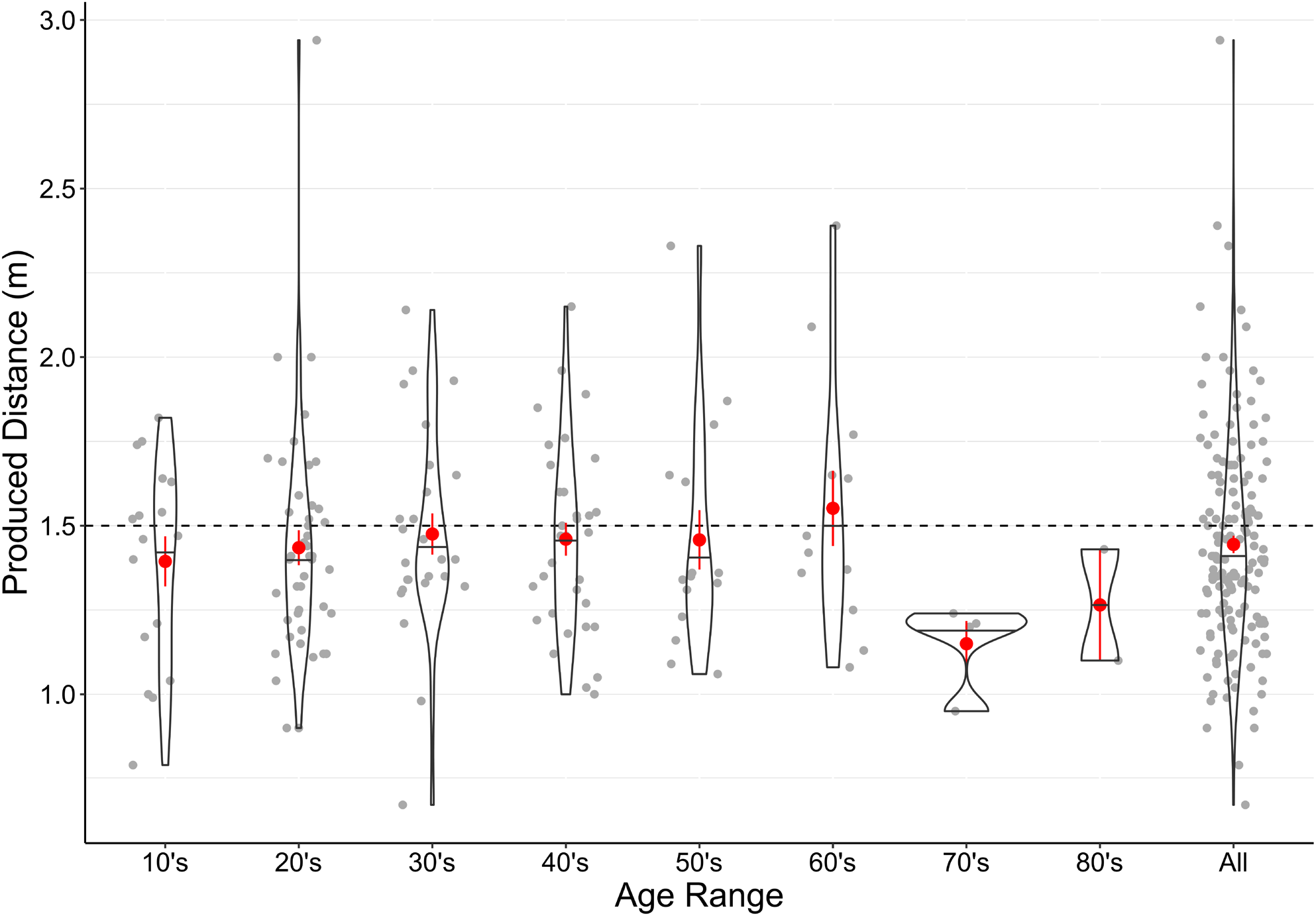
Violin Plots Showing Interpersonal Distances Produced by Participants per Age Range. *Note*. The dashed line denotes the accurate social distance of 1.5 m. Small dots show individual responses made by each participant. They are horizontally jittered within each age range for presentation purposes only—that is, jittering is to reduce overlap of the dots, and the horizontal location of a dot is not to signify each participant’s exact age. For each age range, horizontal bars and large dots indicate median and mean produced distances respectively. Vertical bars represent ±1 standard error of the mean.

In addition to the main analysis described above, two exploratory analyses were carried out for comparing responses between younger and older participants as well as between those who did the task indoors and those who did it outdoors. Due to their exploratory nature, statistical significance was not evaluated in these comparisons. Rather, test results are reported below only for the purpose of describing the data. Non-parametric methods were used for these comparisons because of large discrepancies in the sizes of subsamples that were compared.

A notable pattern in Figure 2 is that participants who were in their seventies and above seemed to leave smaller interpersonal gaps (*M* = 1.188 m, *SD* = 0.145 m) than those who were younger (*M* = 1.455 m, *SD* = 0.320 m). This is interesting because previous studies showed that as compared with younger adults, older adults in this age range judged short distances to be longer (Bian & Andersen, 2013; Norman et al., 2015). A Wilcoxon rank-sum test that examined the difference between responses of older participants (aged 70 and above) and those of younger participants yielded *W* = 189.5, *p* = .018, *d* = 0.841, 95% CI [−0.450, −0.050].

Interpersonal distances produced indoors (*n* = 99, *M* = 1.419 m, *SD* = 0.326 m) and outdoors (*n* = 54, *M* = 1.491 m, *SD* = 0.300 m) were compared because it has been suggested that the same egocentric distance can be verbally reported to be longer in indoor than outdoor environments (Andre & Rogers, 2006; Teghtsoonian & Teghtsoonian, 1970). Figure S1 in Supplementary Material plots the produced distances separately for indoor and outdoor participants. A Wilcoxon rank-sum test comparing indoor and outdoor responses yielded *W* = 2077.5, *p* = .023, *d* = 0.226, 95% CI [−0.210, −0.020].

## Discussion

The present study empirically examined social distancing skills of people in Brisbane, Australia, where the recommended social distance was 1.5 m. Participants were asked to leave exactly 1.5 m from a researcher, and interpersonal distances they produced showed that as a whole, they did this task well—the group mean was just 6 cm shorter than 1.5 m. At first glance, it might look like a reassuring outcome, suggesting that people are able to do social distancing only by being told to keep a certain distance. However, the good performance at the group level does not necessarily provide this assurance: The accurate (but slightly underproducing) group mean actually suggests that many people— more than 50% of the population, if normally distributed—can underproduce the distance when they rely on their space perception. The large inter-individual variability observed in the current data makes this perspective particularly important because the population may form a wide distribution 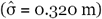, making it relatively common to find individuals who exhibit notable underproduction of interpersonal distances despite their intent to maintain appropriate separation from others.

From a theoretical point of view, the current results were unexpected. It was expected that participants would underperceive egocentric distances to a researcher, which in turn should make them overproduce the requested interpersonal distance. Given the large and robust underperception effects reported in previous studies, it is unlikely that the present study failed to find the predicted pattern due to a type II error (see Supplementary Material for details). Thus, these results call for alternative explanations as to why people on average could produce a distance between themselves and others with accuracy (or with a slight tendency toward underproduction).

As a task of space perception, a notable aspect of the present study is that observers (participants) moved their position to adjust the distance to a stationary target (a researcher). On the other hand, in previous studies that showed underperception of egocentric distance, observers did not move and judged distances to stationary targets (Andre & Rogers, 2006; Bian & Andersen, 2013; Foley et al., 2004; Yamamoto et al., 2014). Thus, as compared with the observers in the previous studies, it is possible that participants in the present study acquired additional cues for distance perception from body movements (e.g., vestibular and proprioceptive cues that inform about the distance walked; Mittelstaedt & Mittelstaedt, 2001). Considering that accurate performance in distance judgement tasks is often obtained when observers indicate to-be-judged distance via bodily action (Loomis & Philbeck, 2008), the availability of the body-based cues could be a major factor for the present participants’ accurate production of the interpersonal distance.

Additionally, potentially insightful observations were noted, where shorter interpersonal distances were produced by (a) older participants (age 70 and above) than younger participants, and (b) participants who did the task indoors than those who performed it outdoors. It has been shown that adults in this age range give greater estimates of visually specified distances as compared with younger adults (Bian & Andersen, 2013; Norman et al., 2015). Thus, relative to the younger participants, the older participants might have overevaluated the interpersonal gaps they left, resulting in their standing closer to a researcher. A similar bias could have caused the indoor versus outdoor difference, as there is some evidence that the same egocentric distance can appear to be longer indoors than outdoors (Andre & Rogers, 2006; Teghtsoonian & Teghtsoonian, 1970). Regardless of underlying mechanisms, these trends may be noteworthy because those who would benefit more from social distancing could have done less of it—that is, older adults are at higher risks of developing serious illness from COVID-19 (Bonanad et al., 2020), and airborne transmission of SARS-CoV-2 is likely to be more prevalent indoors than outdoors (Bulfone et al., 2021). At this stage, given that these trends were derived from exploratory observations, they should be interpreted with caution. They provide important hypotheses that should be investigated in the future.

The present findings carry clear implications for how social distancing recommendations should be provided. On a positive note, they lend empirical support to the way these recommendations are currently given: Just by using space perception and verbal information about a to-be-kept distance, people on average are able to keep this distance with reasonable accuracy. However, they also suggest that under the current recommendations, a non-negligible portion of the population will not leave a sufficient person-to-person gap, even when these people explicitly attempt to do proper social distancing. Therefore, it may be useful for official health advice to include some measures that offset this underproducing tendency. For example, it can deliberately overstate a required interpersonal distance so that the majority of the population will achieve the desirable degree of separation despite some people’s propensity to come too close to others. Using the methodology employed in the present study, the extent to which this overstatement should be done can be empirically specified in future research.

## Data Availability

Original data collected in this study are available on Open Science Framework at https://osf.io/zs7jv/.

## Acknowledgements

The authors thank Nicole Bradshaw, Thessa Canzon, Alani Hope, Stefanie Offen, and Yi Wang for their assistance with data collection.

