## Supplementary text for "How Well Do We Do Social Distancing?"

### A Priori Power Analysis for Computing the Required Sample Size

To examine whether interpersonal distances produced by participants were statistically different from the required social distance (set at 1.5 m in the present study), a two-tailed one-sample *t*-test was planned. An expected effect size was calculated from previous data in which 12 healthy adults aged 30–58 years verbally estimated the distance to a target placed at 1.5 m from them (Yamamoto et al., 2014). Each of these adults gave one estimate of the target distance. On average, they underestimated the distance by 83% ($M$ = 1.24 m, $SD$ = 0.26 m), which led to the conjecture that the egocentric distance of 1.8 m would appear to be 1.5 m. Similar degrees of underestimation were also observed in other studies (Andre & Rogers, 2006; Foley et al., 2004). Thus, in the present study, it was predicted that when participants attempted to stay 1.5-m away from another person, they would result in leaving 1.8 m. Using the standard deviation of verbal estimates given by the 12 adults in the Yamamoto et al. study, the effect size in the present study was estimated to be as large as $d = (1.8 - 1.5) / 0.26 = 1.15$. At $\alpha = .05$ and $1 - \beta = .80$, the planned *t*-test would require nine observations to detect an effect of this size (computed by the G*Power software, version 3.1.9.6; Faul et al., 2007).
