## Supplementary Figure S1 for "How Well Do We Do Social Distancing?"

*Violin Plots Showing Interpersonal Distances Produced Indoors and Outdoors*

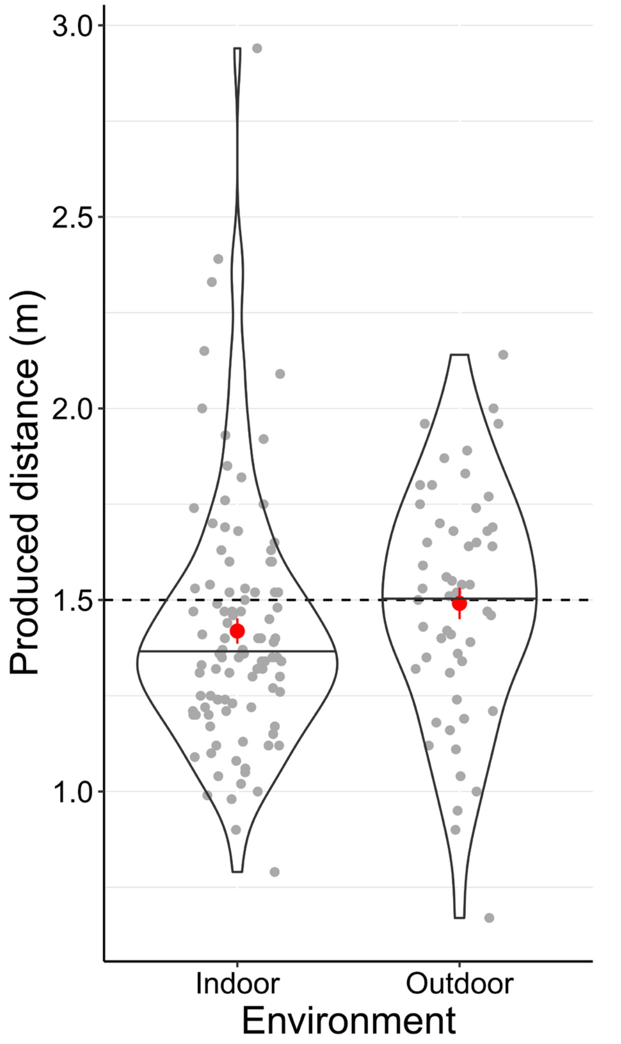


*Note*. The dashed line denotes the accurate social distance of 1.5 m. Small dots show individual responses made by each participant. They are horizontally jittered within each environment to reduce overlap of the dots. For each environment, horizontal bars and large dots indicate median and mean produced distances respectively. Vertical bars represent ±1 standard error of the mean.
